# Dynamics of coagulopathy in patients with different COVID-19 severity

**DOI:** 10.1101/2020.07.02.20145284

**Authors:** Anna Kalinskaya, Oleg Dukhin, Ivan Molodtsov, Alexandra Maltseva, Denis Sokorev, Antonina Elizarova, Olga Sapozhnikova, Ksenia Glebova, Daria Stonogina, Soslan Shakhidzhanov, Evgeniy Nikonov, Alexey Mazus, Ilia Spiridonov, Fazly Ataullakhanov, Leonid Margolis, Alexander Shpektor, Elena Vasilieva

## Abstract

With the progress of COVID-19 studies, it became evident that SARS-CoV-2 infection is often associated with thrombotic complications. The goal of our present study was to evaluate which component of clot formation process including endothelial function, platelets aggregation and plasma coagulation, as well as endogenous fibrinolysis in patients with COVID-19 correlates with the severity of the disease. We prospectively included 58 patients with COVID-19 and 47 healthy volunteers as a control group that we recruited before the pandemic started. It turns out that plasma coagulation with subsequent platelet aggregation, but not endothelial function, correlates with the severity of the COVID-19. IL-6 blockade may play a beneficial role in COVID-19 induced coagulopathy.

## Introduction

With the progress of COVID-19 studies, it became evident that SARS-CoV-2 infection is often associated with thrombotic complications. These complications were first reported at the end of January 2020^1^ and quickly followed by numerous confirming reports on the high prevalence of pulmonary embolism, venous and arterial thrombosis in patients with COVID-19^2^.

There were several brief reports on direct viral infection of the endothelium^3,4^ that in principle can trigger thrombosis. Endothelial damage can activate platelet aggregation via the enhanced release of von Willebrand factor (vWF) on one hand and plasma coagulation via tissue factor release on the other hand^5,6^. Also, there were reports on the hyperproduction of different proinflammatory cytokines (IL-6, TNF-α) that may activate the plasma coagulation process as well^7^. But the exact contribution of the different components of clot formation to the severity of the disease remains unclear.

The goal of our present study was to evaluate which component of clot formation process including endothelial function, platelets aggregation and plasma coagulation, as well as endogenous fibrinolysis in patients with COVID-19 correlates with the severity of the disease.

## Methods

### Subjects

The study was performed at Moscow City Clinical Hospital named after I.V. Davydovsky from May through June 2020. The study protocol was developed in accordance with the principles of the Helsinki Declaration and was approved by the local ethics committee.

We prospectively included 58 patients with COVID-19 and 47 healthy volunteers as a control group that we recruited before the pandemic started. Informed written consent was obtained from all study subjects.

#### Inclusion criteria

patients with a high probability of COVID-19 pneumonia according to the results of a computed tomography (CT-scan), confirmed by the results of polymerase chain reaction (PCR); each patient’s condition was moderate or severe (criteria are summarized in Table 1); informed written consent.

**Table №1.** Criteria for Patient Conditions

| Moderate | Severe |
| --- | --- |
| • Body temperature above 37.5 ° C.<br>• Respiratory rate 22-30/min;<br>• Exertional dyspnea;<br>• Visualization data (stage 1 or 2 of COVID-19 pneumonia according to results of CT-scan;<br>• Increased serum C-reactive protein (CRP) level ($\geq 10$ mg/L); | • Respiratory rate $>30/\text{min}$<br>• $\text{SpO}_2 \leq 93\%$<br>• $\text{PaO}_2 / \text{FiO}_2 \leq 300 \text{ mmHg}$<br>• Visualization data (stage 3 or 4 of COVID-19 pneumonia according to results of CT-scan;<br>• Decrease in the level of consciousness, agitation<br>• Arterial blood lactate $> 2 \text{ mmol / L}$<br>• qSOFA $> 2$ points |

#### Exclusion criteria

previous use of anticoagulants; unconfirmed diagnosis of COVID-19; We worked with a group of healthy volunteers without signs of cardiovascular and pulmonary chronic diseases. The control group was assembled from September to December 2019.

### Clinical assessment

All patients underwent chest CT upon admission (and, if necessary, repeatedly in dynamics) and in dynamics, physical examination, standard laboratory tests, including D-dimer, activated partial thromboplastin time (aPTT), prothrombin time (PT), international normalized ratio (INR), fibrinogen, and legs-vein ultrasound. Healthy volunteers underwent a physical examination and a serum blood test.

#### Criteria for Patient Conditions

In addition to the standard clinical examination, on admission (point 1), all patients underwent:

1. Flow-mediated dilation test (FMD-test) to evaluate endothelial function,
2. Optical aggregation, impedance aggregation to evaluate platelets function,
3. Thrombodynamics, rotational thromboelastometry (in NATEM mode) to evaluate plasma coagulation.
4. Thrombodynamics in fibrinolysis mode to evaluate endogenous fibrinolysis.

Subsequently, all studies were repeated on the 3rd day of the patient’s stay in the hospital (point 2) and on the 9th day (point 3).

### FMD test

In order to assess endothelial function, all patients underwent an FMD-test according to standard method^8^. Briefly, the essence of the method consists in sonographic registration of the change of the diameter of the brachial artery after its five-minute clamping. Mechanical clamping of the artery leads to the release of NO which is a powerful vasodilator. The low values of this study are described for diseases such as myocardial infarction, Takotsubo syndrome, etc^9,10^. The FMD test is widely known as an independent predictor of adverse cardiovascular events^11^.

### Blood sampling

The blood samples were obtained prior to the first anticoagulant administration. A 21-gauge needle was used with minimal stasis in order to avoid iatrogenic induction of platelet aggregation, and peripheral venous blood was drawn in the amount of 4.5 ml into a tube containing 0.105 M buffered sodium citrate anticoagulant. At points 2 and 3, blood samples were collected between 6:00 and 6:30 a.m. or between 5:30 and 6:00 p.m. (12 hours after the last injection of the anticoagulant). In cases of continuous infusion of unfractionated heparin, blood sampling was carried out without a clear time reference. The delay from the moment of blood sampling until the start of the study was no more than 15 minutes.

The whole blood was used for impedance aggregometry and rotational thromboelastometry. Platelet-rich plasma was used for optical aggregometry. Platelet-poor plasma was used for the Thrombodynamics study.

### Optical Aggregometry

The study was conducted on an ALAT-2 Analyzer (Biola)^12^. For analysis, an indicator of spontaneous platelet aggregation (arbitrary units) was used.

### Impedance Aggregometry

To assess impedance aggregometry, we used the Multiplate analyzer ^13,14^. Arachidonic acid, adenosine diphosphate (ADP), thrombin receptor-activated peptide (TRAP) and ristocetin (RISTO) were used for platelet’s activation. The test time was 6 minutes. Platelet aggregation was assessed from the area under the curve (AUC).

### Rotational thromboelastometry

We carried out the study using ROTEM (Roche). The study was conducted in NATEM mode. For the analysis, the following indicators were used: clotting time (CT, sec), clot formation time (CFT, sec), thrombus amplitude at different time sections of the study (A5-A30, mm), angle α (α, °), thrombus lysis index at different time sections of the study (Li30%, Li45%, Li60%), the time of lysis onset (LOT, s), and maximum thrombus growth rate (MaxV mm/min).

### Thrombodynamics

The study was performed on a T-2 Thrombodynamics Analyzer (Hemacore). The study was carried out according to a standard technique^15^. The following parameters of clot growth were used: clot growth rate (V, μm /min), initial and stationary clot growth rate (Vi, μm /min; Vst, μm/min), Lag-time (Tlag, min), clot size (CS, μm), clot density (D, arb units), spontaneous clots formation time (Tsp, min). A standard activator with urokinase was added to induce thrombus lysis. As the parameters of lysis, the time of lysis onset (LOT, min), the rate of lysis progression (LP, %/min), the thrombus lysis time (CLT, min), and the expected clot lysis time (LTE, min) were evaluated.

### Combination of methods

Thromboelastometry in NATEM mode allows to measure the parameters of thrombus formation in whole blood without using any coagulation activators, but this method has limitations in fibrinolysis evaluation. The main advantage of thrombodynamics is the modeling of damage to the vascular wall, which leads to exposure of tissue factor (TF) and subsequent activation of coagulation. Moreover, the modification of this test by adding the small amounts of urokinase allows a more accurate assessment of the state of fibrinolysis. The combination of these techniques with aggregometry (optical and impedance) allows to obtain the maximum information about plasma and platelet coagulation, endogenous fibrinolysis.

### Influence of tocilizumab

We studied the effect of the interleukin-6 receptor blocker tocilizumab on the main parameters of clot formation and fibrinolysis, endothelial function. Among the patient population, 16 patients received tocilizumab (of whom 10 were representatives of the severe subgroup). There were no significant differences in the clinical data; however, patients receiving tocilizumab were characterized by a higher level of inflammation markers (hs-CRP, aspartate aminotransferase (AST), lactate dehydrogenase (LDH)), a lower lymphocyte count, and a shorter PT (see supplemental table №1)

### Statistical analysis

Statistical analysis was performed with the Python 3 programming language using the *numpy, scipy*, and *pandas* packages. The Mann–Whitney U test (two-sided, with continuity correction) was used for comparing distributions of quantitative parameters between independent groups of patients. The Fisher exact test (two-tailed) was used for comparing qualitative parameters between independent groups of patients. The Wilcoxon signed-rank test (two-sided, including zero-differences in the ranking process and splitting the zero rank between positive and negative ones) was performed to assess the changes in the quantitative parameters between different time points for the same patient. The significance level α for *p*-values was set to 0.05. In order to control type I error, FDR *q*-values were calculated independently for each family of statistical comparison using the Benjamin–Hochberg (BH) procedure, and a threshold of 0.1 was set to keep the positive false discovery rate below 10%. The chi-square test of independence of variables was used for comparison of CT-stage distributions between patient subgroups.

## Results

### Clinical data

1. Comparison of the control group and patients with COVID-19. Patients with COVID-19 were significantly more likely to suffer from arterial hypertension than were members of the control group. According to laboratory tests, there were increases in creatinine, total cholesterol, hs-CRP, glucose, and AST in the COVID-19 group (see supplemental table 2).
2. Comparison of patient subgroups. Our study included 58 patients with COVID-19: 39 in the moderate subgroup and 19 in the severe subgroup. There were no significant differences between the subgroups of patients in baseline characteristics (see supplemental table 3). Patients in the severe subgroup were characterized by a more significant increase in markers of systemic inflammation (hs-CRP, procalicitonine (PCT), transaminases) and D-dimer (see supplemental table 4).

During hospitalization, all patients received anticoagulant therapy. In the severe subgroup, 56% of patients received tocilizumab, while 18% of patients in the moderate subgroup received it. We had seven patients (all from the severe subgroup) who required invasive or non-invasive ventilation; two of them died.

Leg-vein ultrasound was performed in 80% of patients. One patient had deep vein thrombosis of legs.

### Flow-mediated dilation test

1. In the majority of patients with COVID-19, values on the FMD-test were predominantly within the reference range; only in 30% of patients impaired endothelial-dependent vasodilatation was observed (FMD < 5%). Patients with COVID-19 were characterized by a slightly reduced value of endothelium-dependent vasodilation compared with the control group, with the mean value remaining in the normal range (FMD, % 9.2 [6.4; 15.1] vs 7.14 [4.2; 11.1], p=0.02). Fig 1.
2. There were no significant differences in the FMD-test results between the patient’s subgroups. The endothelium-dependent vasodilation values varied from 0.89% to 24% independently of the severity of the disease. At point 3 the FMD test increased in both subgroups. Fig 1.

**Figure 1.**
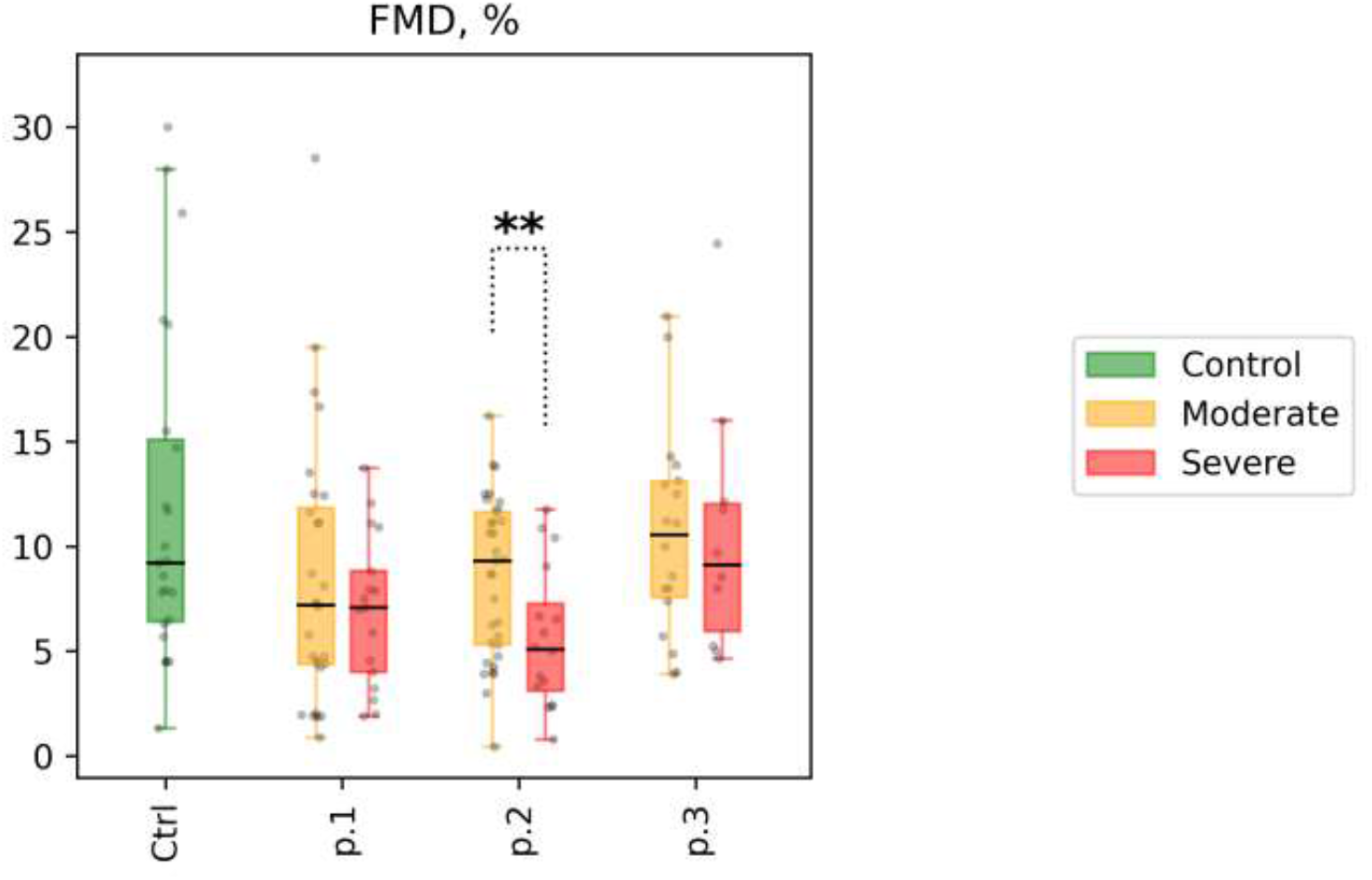
Flow-mediated dilation test (FMD, %) for patient subgroups at time points 1, 2, and 3 (shown as p.1, p.2, and p.3 at the x-axis, respectively). Distribution of values in the control group is shown for reference (shown as Ctrl at the x-axis). Mann-Whitney p-values are shown for comparisons of distributions between the moderate and the severe group at different time points: * p-value<0.05, q-value>0.1; ** p-value<0.05, q-value<0.1.

### Platelet aggregation

1. Patients with COVID-19 showed lower values of ADP-induced platelet aggregation (AUC ADP, 43.9 [34.0; 50.0] vs 48.5 [42.0; 53.3], p=0.04) and thrombin-induced platelet aggregation (AUC Thrombin, 60.0 [41.0; 71.0] vs 67.5 [60.8; 86.3], p=0.05; the comparison with the control group remains significant after correction for multiple comparisons, with FDR<0.1). No difference was observed for platelet aggregation induced by arachidonic acid (AUC Asa; see supplemental table 5, Fig 2).
2. The indicators of spontaneous platelet aggregation in patients with COVID-19 in both the moderate and the severe subgroups were within the reference values, but closer to the lower boundary of the reference interval. Significant differences in spontaneous platelet aggregation were not observed either at point 1 or in subsequent measurements (Figs. 2 and 3).

**Figure 2.**
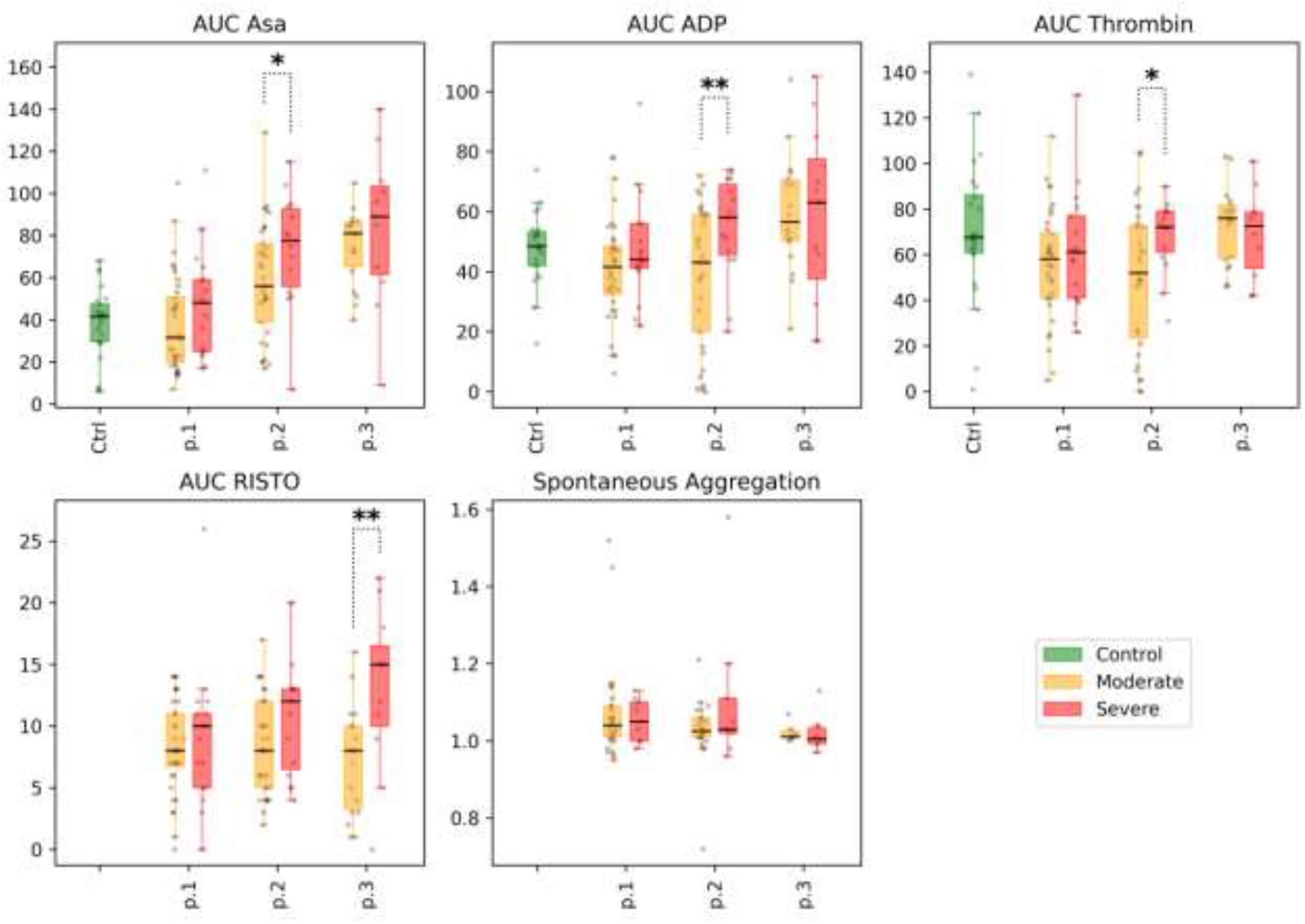
Impedance Aggregation and Optical Aggregation: selected parameters are arachidonic acid-induced aggregation (AUC ASA), %; ADP-induced aggregation (AUC ADP), %; thrombin-induced aggregation (AUC Thrombin), %; ristocetin-induced aggregation (AUC RISTO), %; spontaneous aggregation, arb. units.) for the different subgroups of patients at time points 1, 2, and 3 (labeled p.1, p.2,and p.3 at the x-axis, respectively). Distribution of values in the control group is shown for reference (labeled Ctrl at the x-axis, for each of the parameters). Mann-Whitney p-value are shown for comparisons of distributions between the moderate and severe groups at different time points: * p-value<0.05, q-value>0.1; ** p-value<0.05, q-value<0.1.

**Figure 3.**
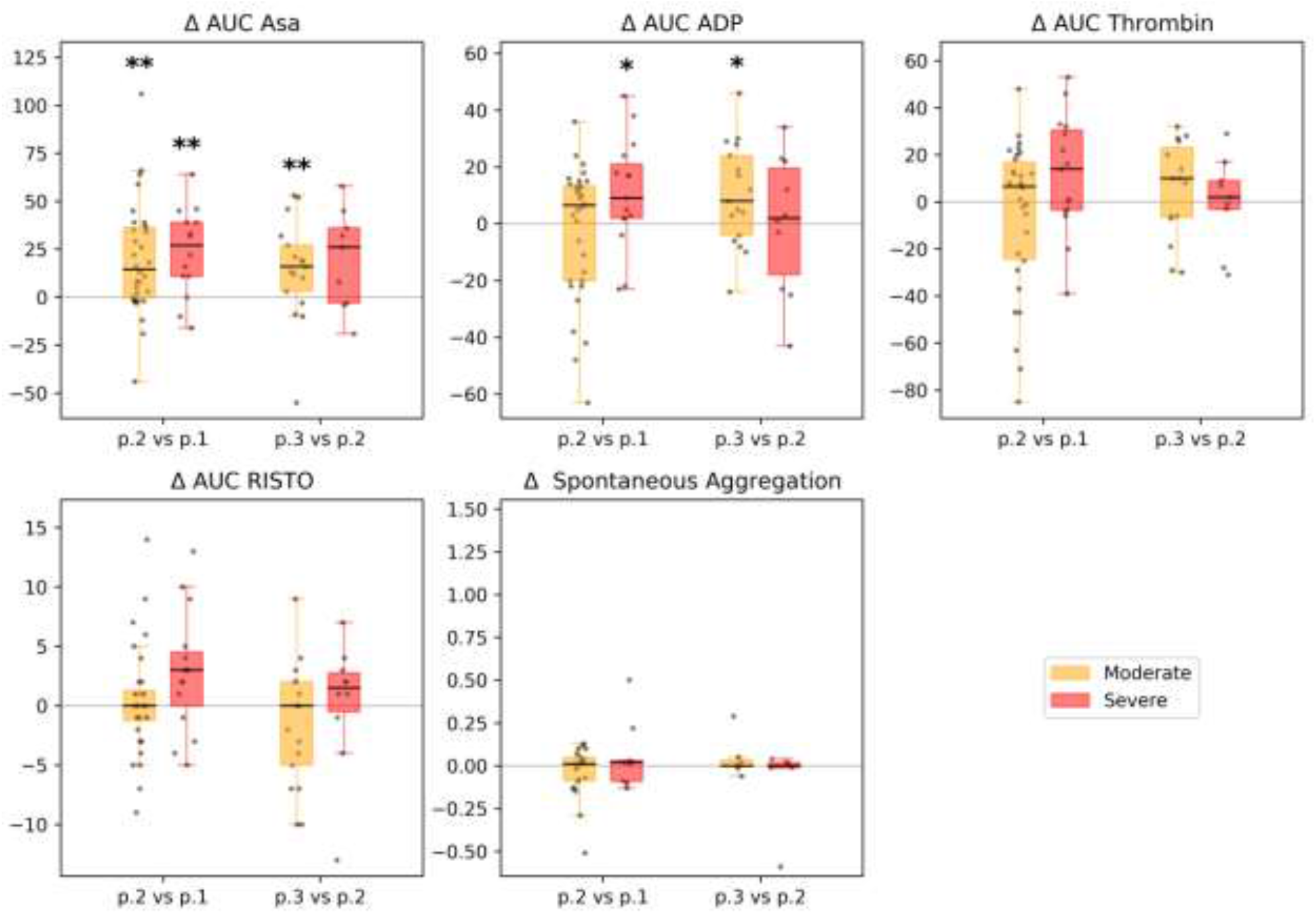
Impedance Aggregation and Optical Aggregation: selected parameters (arachidonic acid-induced aggregation (AUC ASA), %; ADP-induced aggregation (AUC ADP), %; thrombin-induced aggregation (AUC Thrombin), %; ristocetin-induced aggregation (AUC RISTO), %; spontaneous aggregation, arb. units.) between time points in different subgroups of patients. Each patient’s individual changes in parameter levels at different time points were used as a basis for box-and-whiskers plot. Points of comparison are shown at the x-axis as ‘p.2 vs p.1’ for differences between point 2 and point 1, and as ‘p.3 vs p.2’ for differences between point 3 and point 2. The Wilcoxon signed-rank test p-values for comparison between time points are shown: * p-value<0.05, q-value>0.1; ** p-value<0.05, q-value<0.1.

There were no significant differences between the subgroups studied at point 1 in platelet aggregation induced by arachidonic acid, ADP, thrombin, or ristocetin. The indices of induced aggregation were at the lower level of the reference values. In dynamics, an increase in the indicators of aggregation induced by arachidonic acid was observed in both subgroups. Increases in ADP, thrombin, and ristocetin aggregation were more pronounced in the severe subgroup (however, these indicators remained within the reference values); see Figs. 2 and 3. Full details are presented in Supplemental Table 6.

### Rotational thromboelastometry

1). Patients with COVID-19 were characterized by faster growth of the thrombus (CT, sec, 638.5 [481.0; 848.5] vs. 832.0 [709.0; 937.0], *p*<0.01) and its larger size (A30, mm, 59.0 [53.0; 64.0] vs. 53.0 [47.0; 56.5], *p*<0.01) at different time sections of the experiment, as well as an earlier onset of clot lysis (LOT, sec, 5 108 [4 655.5; 5 214.5] vs. 5 674.0 [5 090.0; 8 093.0], *p*=0.05) compared with the control group (see Supplemental Table 7 and Fig.4).

2). There was no significant difference between the patient subgroups in thrombus size and thrombus growth rate in different time sections of the study. But there was a significant decrease in the size and growth rate of the thrombus in the moderate subgroup at point 2 compared with point 1. In the severe subgroup, we observed only a trend. At point 3 we noted a slight elevation of the thrombus growth rate in the moderate subgroup, which can be explained by the reduction of the anticoagulation therapy before the discharge (Figs. 4 and 5). Full details are presented in Supplemental Table 8.

**Figure 4.**
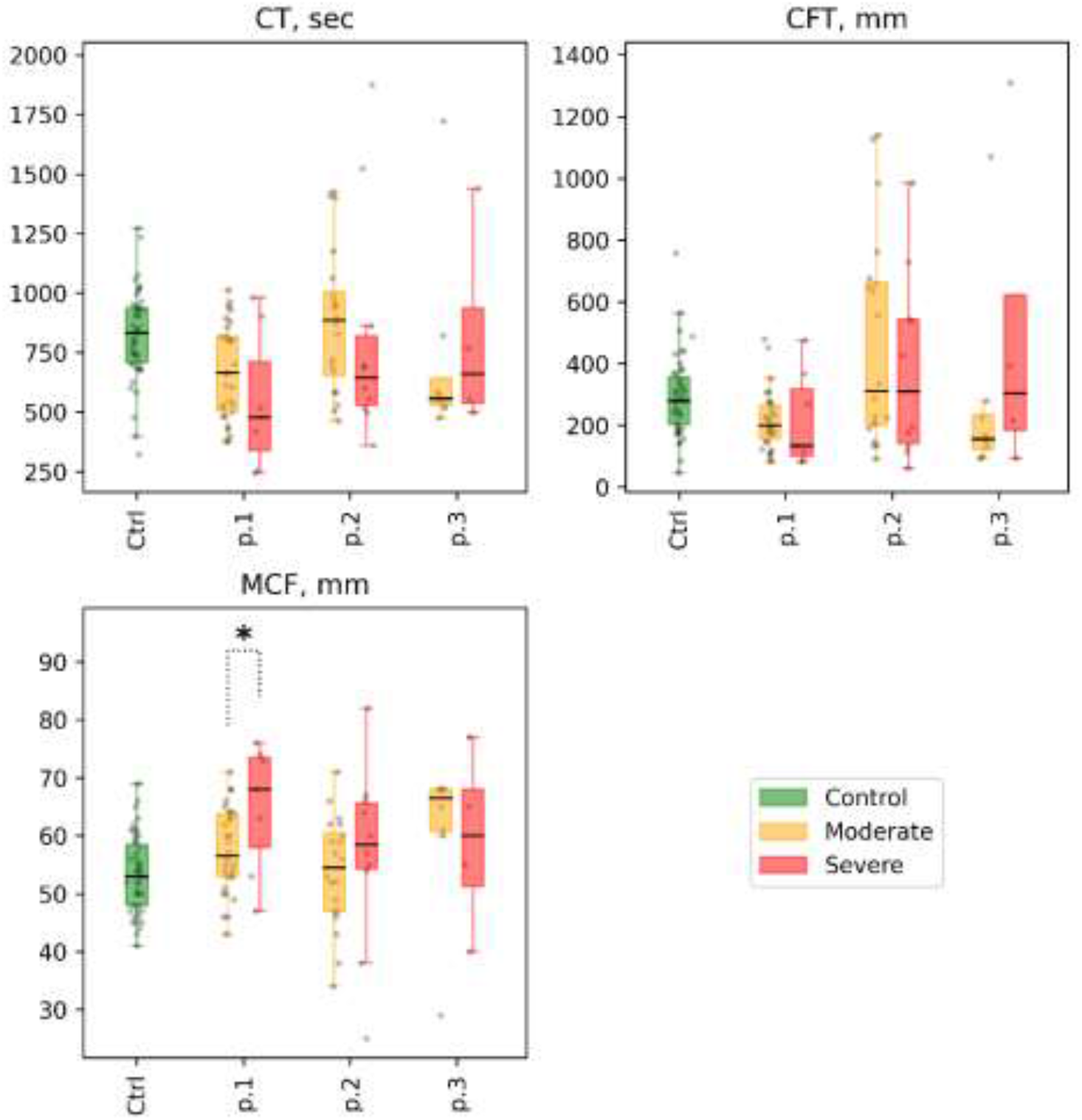
ROTEM (in NATEM mode): selected parameters are clotting time (CT), sec; clot formation time (CFT), sec; and maximum clot firmness (MCF), mm, for different subgroups of patients at time points 1, 2, and 3 (labeled p.1, p.2, and p.3 at the x-axis, for each parameter).. Distribution of values in the control group is shown for reference as Ctrl at the x-axis, for each parameter). Mann-Whitney p-value are shown for comparisons of distributions between moderate and severe groups at different time points: * p-value<0.05, q-value>0.1; ** p-value<0.05, q-value<0.1.

**Figure 5.**
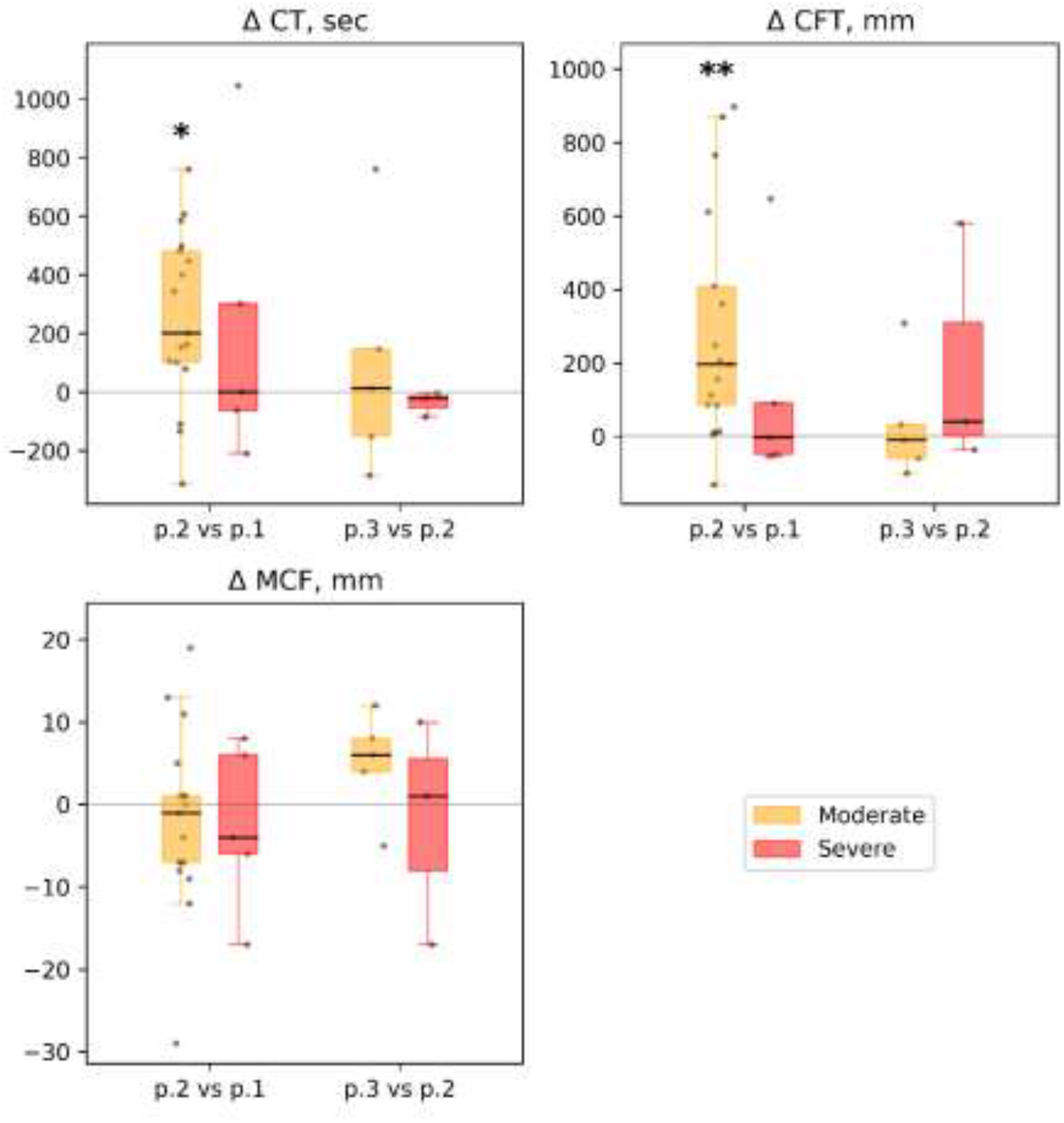
ROTEM (in NATEM mode): dynamics of selected parameters between time points in different subgroups of patients: clotting time (CT), sec; clot formation time (CFT), sec; maximum clot firmness (MCF), mm. Each patient’s individual changes in parameter levels at different time points were used as a basis for box-and-whiskers plot. Points of comparison are labeled at the x-axis as ‘p.2 vs. p.1’ for differences between point 2 and point 1 and as ‘p.3 vs. p.2’ for differences between point 3 and point 2. The Wilcoxon signed-rank test p-values for comparison between time points are shown: * p-value<0.05, q-value>0.1; ** p-value<0.05, q-value<0.1.

### Thrombodynamics

1. Patients with COVID-19 had greater optical density of the clot (D, arb units, 29 425.0 [26 724.0; 31 945.0] vs. 26 913.0 [23 858.5; 30 786.8], *p*=0.01) as well as an earlier appearance of spontaneous clots in plasma (Tsp, min, 29.7 [19.0; 54.2] vs 55.5 [44.4; 85.7], *p*<0.01) compared with the control group. We did not find any significant differences in the average and initial clot growth rates (see Supplemental Table 9 and Fig 6).
2. A higher optical density of the fibrin clot was found in the severe subgroup of patients than in the moderate subgroup (D, arb units 28 212.0 [26 229.0; 29 939.0] vs. 31 630.0 [29 690,0; 35 770.0], *p*=0.02), but it did not remain significant after correction for multiple comparisons with FDR > 0.1. We noted a moderate positive correlation between the optical density of the clot (D, arb units) and fibrinogen concentration in both subgroups (Spearman R=0.55; *p*<0.01). No significant differences were found in the mean and initial clot growth rates. The dynamics showed a decrease in the rate of growth of the clot (size and density) in both subgroups of patients. However, this effect was more pronounced among patients with moderate severity of the disease (Figs. 6 and 7). Full details are presented in Supplemental Table 10.

**Figure 6.**
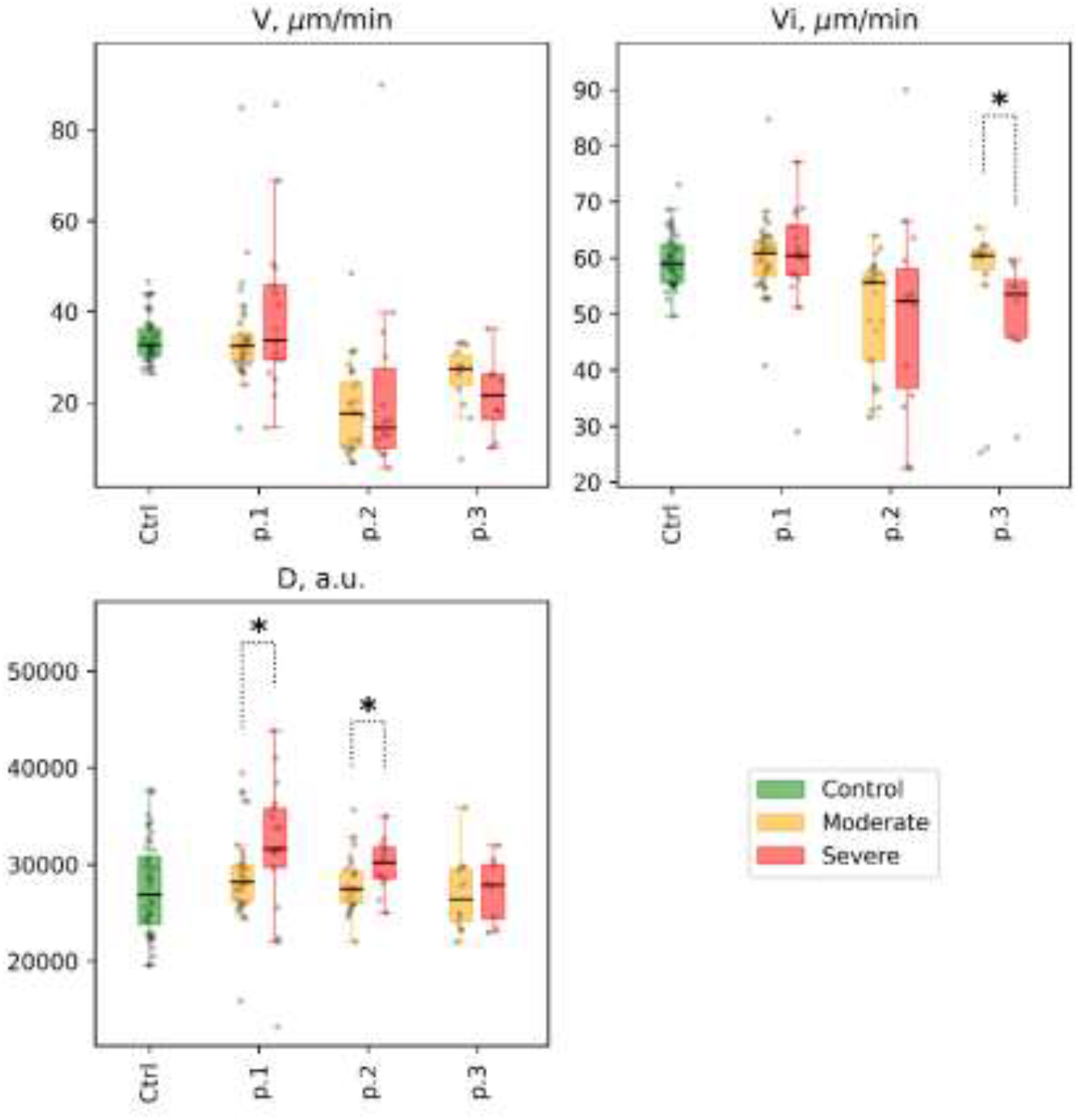
Thrombodynamics. Selected parameters are shown for different patient subgroups at time points 1, 2, and 3 (labeled as p.1, p.2, and p.3, respectively), at the x-axis: clot growth rate (V), μm/min; initial clot growth rate (Vi), μm/min; clot density (D), arb units). Distribution of values in the control group is shown for reference (labeled as Ctrl at the x-axis, for each parameter). Mann-Whitney p-value are shown for comparisons of distributions between moderate and severe groups at different time points: * p-value<0.05, q-value>0.1; ** p-value<0.05, q-value<0.1

**Figure 7.**
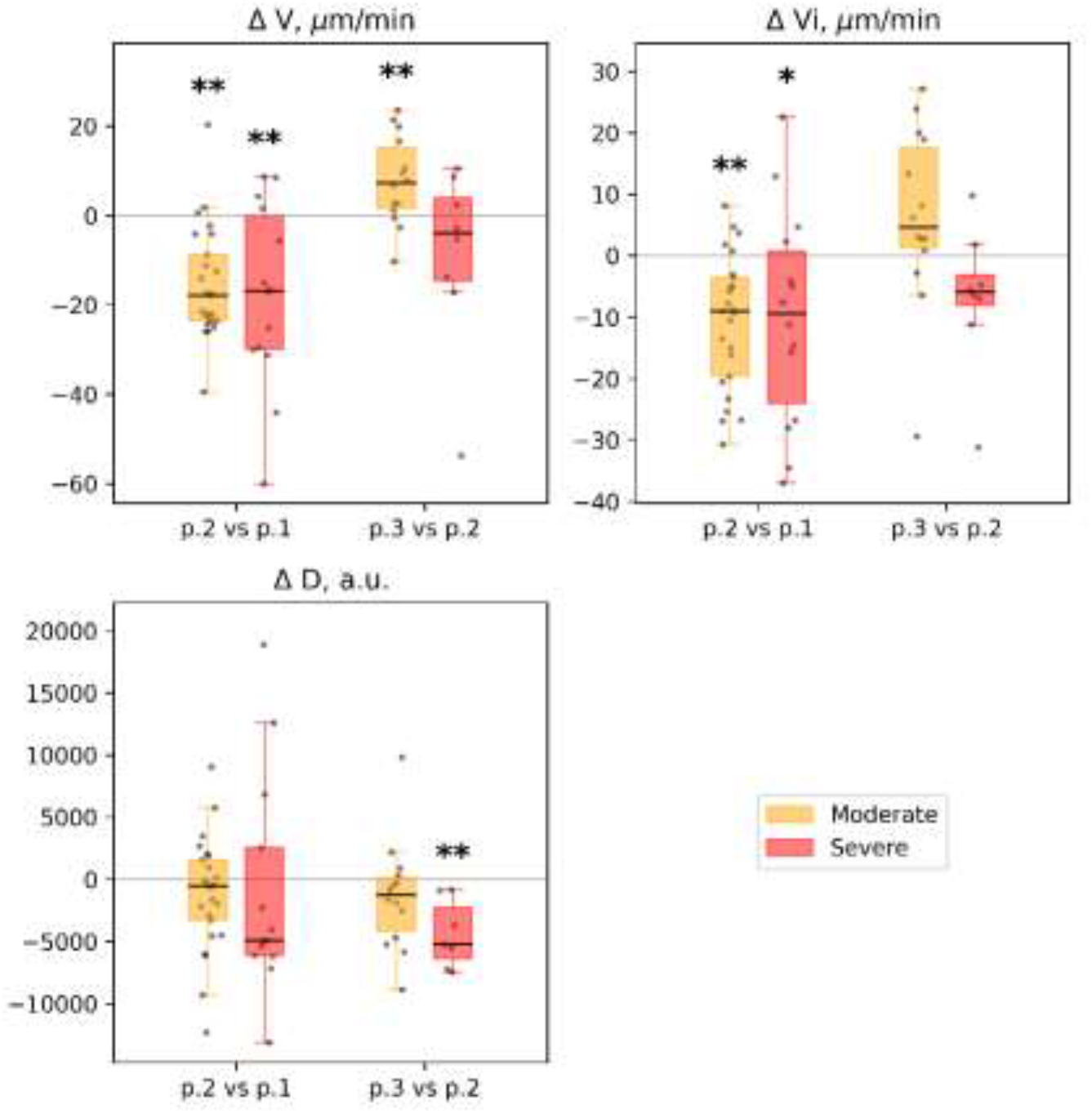
Thrombodynamics: dynamics of selected parameters recorded between time points in different groups of patients: clot growth rate (V), μm/min; initial clot growth rate (Vi), μm/min; clot density (D), arb units. Each patient’s individual changes in parameter levels at different time points were used as a basis for box-and-whiskers plot. Points of comparison are labeled at the x-axis as ‘p.2 vs. p.1’ for differences between point 2 and point 1, and ‘p.3 vs. p.2’ for differences between point 3 and point 2. The Wilcoxon signed-rank test p-values for comparison between time points are shown: * p-value<0.05, q-value>0.1; ** p-value<0.05, q-value<0.1.

### Fibrinolysis

1. Patients with COVID-19 were characterized by an earlier onset of clot lysis (LOT, min, 30.5 [26.8; 38.5] vs. 63.3 [42.6; 80.7], *p*<0.01) as well as its faster and more effective lysis (LP,%/min, 5.2 [3.1; 8.1] vs 2.0 [1.4; 3.4], *p*<0.01). (See Supplemental Table 11 and Fig 8)
2. We found no significant differences in the parameters of fibrinolysis between patient subgroups both at point 1 and in subsequent measurements (see Supplemental Table 12 and Figs. 8 and 9).

**Figure 8.**
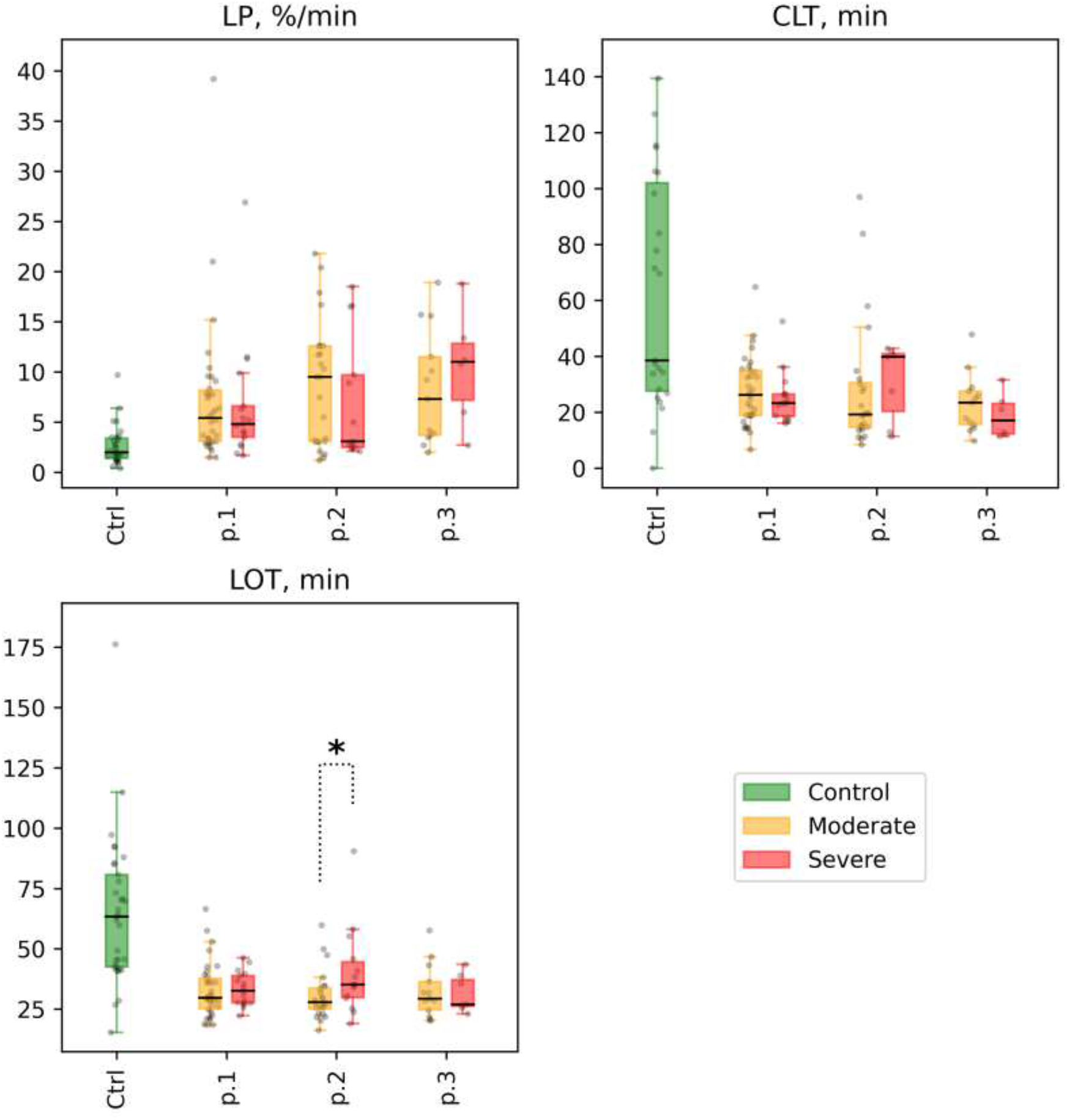
Fibrinolysis. Selected parameters for different groups of patients at time points 1, 2, and 3 (labeled as p.1, p.2, and p.3 at the x-axis, for each parameter): lysis progression (LP), %/min; clot lysis time (CLT), min; lysis onset time (LOT), min. Distribution of values in the control group is shown for reference (labeled as Ctrl at the x-axis, respectively). Mann-Whitney p-values are shown for comparisons of distributions between the moderate and severe groups at different time points: * p-value<0.05, q-value>0.1; ** p-value<0.05, q-value<0.1.

**Figure 9.**
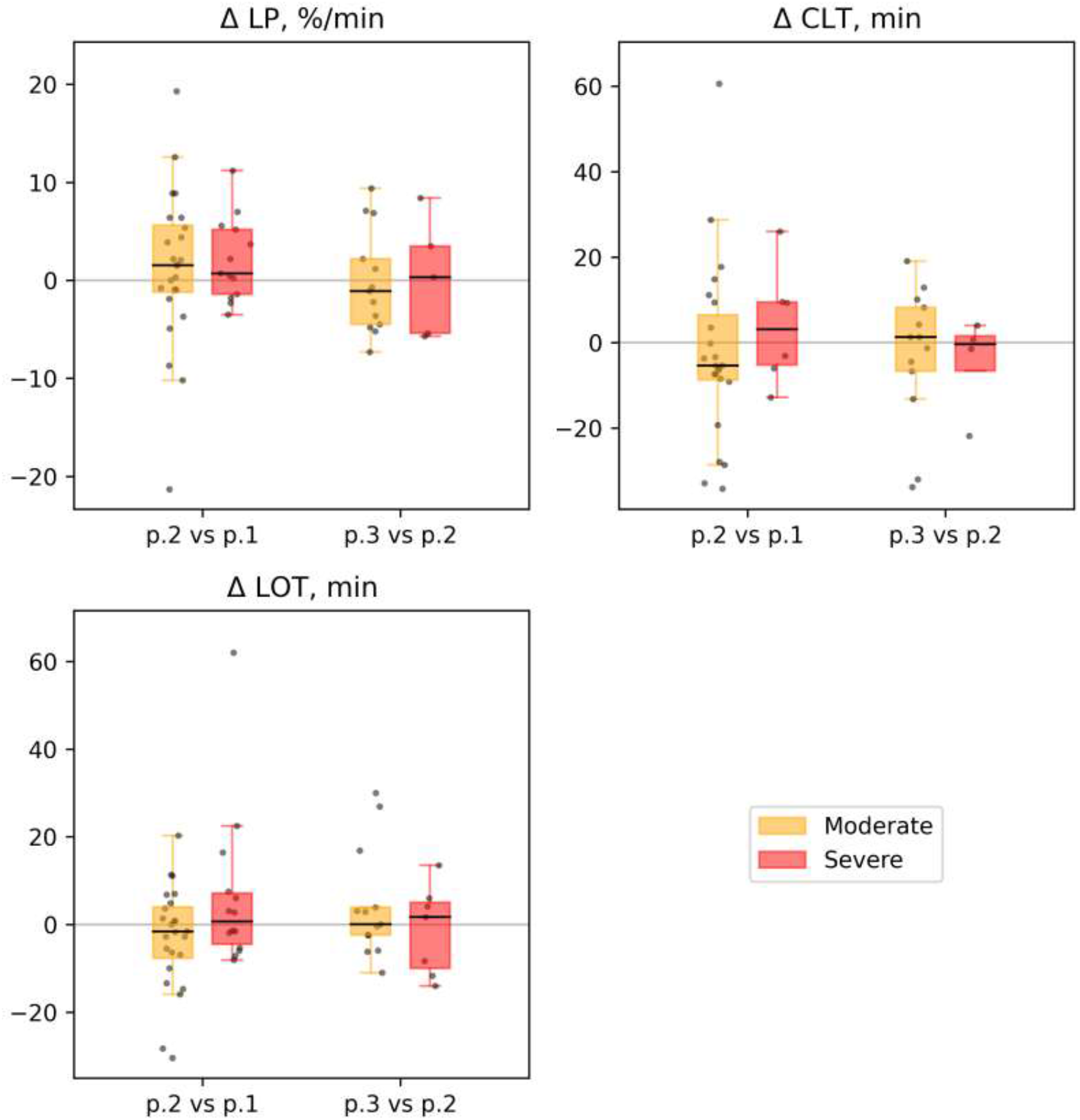
Fibrinolysis. The dynamics of selected parameters for different groups of patients at time points 1, 2, and 3 (labeled as p.1, p.2 and, p.3 at the x-axis, for each parameter): Lysis progression (LP), %/min; clot lysis time (CLT), min; lysis onset time (LOT), min. Each patient’s individual changes in parameter levels at different time points were used as a basis for box-and-whiskers plot. Points of comparison are labeled at the x-axis as ‘p.2 vs. p.1’ for differences between point 2 and point 1 and as ‘p.3 vs. p.2’ for differences between point 3 and point 2. The Wilcoxon signed-rank test p-values for comparison between time points are shown: * p-value<0.05, q-value>0.1; ** p-value<0.05, q-value<0.1.

### D-dimer

Patients in severe subgroup were characterized by a higher level of D-dimer compared to the moderate severity subgroup (D-dimer, ng/ml 864.0 [454.0; 1 614.0] vs 344.0[209.5; 793.0], p<0.01).

### Influence of tocilizumab on the clot formation and lysis processes

We studied the effects of tocilizumab on the clot formation and lysis processes. Because of the low number of patients treated with tocilizumab (*n*=16), none of effects mentioned below demonstrated FDR<0.1 after correction for multiple comparisons. Still, patients after the tocilizumab infusion were characterized by smaller clot size (A25, mm, 51.0 [37.5;57.0] vs. 65.0 [61.0; 68.0], *p*-value<0.05) and prolonged clot formation time (CFT, min, 393.0 [304.5; 851.0] vs 146.0 [98.0; 224.0], *p*-value<0.05) compared with tocilizumab non-receivers.

ADP- and Thrombin-induced platelet aggregation was lower in patients after the infusion of tocilizumab than in tocilizumab non-receivers (ADP 48.5 [24.3; 60.0] vs. 62 [49.0; 73.5], *p*-value<0.05; TRAP 57.0 [44.3; 75.0] vs. 76.5 [61.5; 83.5], *p*-value<0.05).

Tocilizumab receivers had lower parameters of fibrinolysis than non-receivers (LOT, min, 35.4 [34.0; 58.1] vs. 28.0 [25.4; 34.6], *p*-value<0.05; LP, %/min, 2.7 [2.3; 9,1] vs. 9.5 [3.1; 12.6], *p*-value<0.05). There were no significant differences in the results of the FMD-test. (See Supplemental Table 13).

## Discussion

Hypercoagulation is described for various viral infections: H1N1-influenza, Ebola virus, HIV, Hepatitis C, Cytomegalovirus, and others^16–20^.

COVID-19 is particularly often complicated by coagulopathy, which can lead to DIC and various thromboembolic complications^21–24^. Nahum et al. reported high prevalence of deep vein thrombosis among critically ill patients (about 65-79%) with COVID-19^25^. Moreover, in post-mortem study of patients died from COVID-19 platelet–fibrin thrombi in small arterial vessels were observed in 87% of cases^26^. In the studies mentioned above patients did not receive routine anticoagulant therapy even in intensive care unit. According to N. Tang et al., heparin administration reduced 28-day mortality among high-risk patients (SIC score> 4 or D-dimer >6-fold of upper limit) compared with patients who did not receive it^27^.

The processes of clot formation consist of the following main components: endothelium transformation, platelet activation, activation of plasma clotting factors, and fibrinolysis. In order to evaluate possible contributions of each of these factors into clot formation in SARS CoV-2 infected patients we evaluated them separately in relation to the severity of the COVID-19.

The possible role of endothelial damage and dysfunction in COVID-19 progression was described previously^5,28–30^ and in principle can be caused by direct endothelium infection with SARS-CoV2 ^3,4^.

Here, we evaluated endothelial function with the FMD-test that reports on the ability of endothelial cells to release NO. Higher NO release correlates with the normal state of endothelial. 70% of patients had FMD-test level in the normal range and even in some patients it was elevated to 24%, that is 2,0-2,5 higher than in average healthy persons (12,0 ± 3,2)^11,31^. We did not observe any significant difference between the severely and moderately ill subgroups.

Despite the fact that the FMD-test has limitations in assessing small-caliber vessels, in our study we did not observe generalized endothelial dysfunction.

In our study the initial depression of platelet aggregation in patients compared with the control group was followed by a significant increase in platelet reactivity during disease progression. This increase was more pronounced in the severely ill patients, in agreement with literature data^6^. We demonstrated for the first time the deferred mode of this activation.

Nevertheless, platelet aggregation values in patient subgroups ranged between lower and upper limits reported for normal individuals.

In contrast, we found out that plasma coagulation activation was much more active in COVID-19 patients than in the control group. Furthermore, the level of activation correlated with the severity of the disease in particular during disease progression. It is important that this activation was evaluated despite the 100% usage of anticoagulants in our patients. Optimal anticoagulation therapy, especially in severe patients, needs further study.

Also, we found that the process of endogenous fibrinolysis was significantly more active in COVID-19 patients compared to control group. However, we did not find any differences in process of endogenous fibrinolysis between the severely and moderately ill subgroups of patients, with the exception of elevated levels of D-dimer (in particular, among severely ill patients), like many other authors did ^22,32^.

Thus, study revealed the leading role of plasma coagulation, rather than endothelial dysfunction in COVID-19 induced coagulopathy. It seems that the damage of lung and sometimes other tissues, including myocardium, together with macrophages activation and particularly cytokines release lead to massive release of tissue factor that triggers plasma coagulation.

We attempted to evaluate whether suppression of cytokines in COVID-19 patients affected blood clotting. Several publications reported on tocilizumab, an efficient inhibitor of IL-6 receptor suppress the “cytokine storm” and is beneficial for SARS CoV-2 patients at certain stages of the disease^33,34^.In our assays we observed a decrease of clot formation and fibrinolysis among tocilizumab-treated patients. Although this decrease was significant when evaluated as a single parameter, the difference from samples from tocilizumab did not reach statistical significance after multiple comparison correction. A larger cohort of treated patients is needed to confirm the effect cytokine suppression on clot formation.

## Conclusion

Plasma coagulation with subsequent platelet aggregation, but not endothelial function, correlates with the severity of the disease. IL-6 blockade may play a beneficial role in COVID-19 induced coagulopathy.

## Data Availability

All data collected during the study are presented in the manuscript. All additional information can be provided with the reasonable response to corresponding author.

